# Advanced Deep Learning Architecture for the Early and Accurate Detection of Autism Spectrum Disorder Using Neuroimaging

**DOI:** 10.1101/2025.08.30.25333188

**Authors:** Nazeela Fatima, Nargis Bibi, Amad Ud Din

## Abstract

Autism Spectrum Disorder (ASD) is a neurological condition that affects the brain, leading to challenges in speech, communication, social interaction, repetitive behaviors, and motor skills. This research aims to develop a deep learning based model for the accurate diagnosis and classification of autistic symptoms in children, thereby benefiting both patients and their families. Existing literature indicates that classification methods typically analyze region based summaries of Functional Magnetic Resonance Imaging (fMRI). However, few studies have explored the diagnosis of ASD using brain imaging. The complexity and heterogeneity of biomedical data modeling for big data analysis related to ASD remain unclear. In the present study, the Autism Brain Imaging Data Exchange 1 (ABIDE-1) dataset was utilized, comprising 1,112 participants, including 539 individuals with ASD and 573 controls from 17 different sites. The dataset, originally in NIfTI format, required conversion to a computer-readable extension. For ASD classification, the researcher proposed and implemented a VGG20 architecture. This deep learning VGG20 model was applied to neuroimages to distinguish ASD from the non ASD cases. Four evaluation metrics were employed which are recall, precision, F1-score, and accuracy. Experimental results indicated that the proposed model achieved an accuracy of 61%. Prior to this work, machine learning algorithms had been applied to the ABIDE-1 dataset, but deep learning techniques had not been extensively utilized for this dataset and the methods implied in this study as this research is conducted to facilitate the early diagnosis of ASD.

## Introduction

Autism spectrum disorder (ASD) is a neurological condition that affects the brain, leading to difficulties in speech, communication, social interaction, repetitive behaviors, and cognitive abilities. This disorder impacts multiple areas of the brain. The main causes of autism spectrum disorder are genetic and environmental factors. Globally, autism affects one in every 70 children. In 2018, the United States had one of the highest prevalence rates, with autism spectrum disorder estimated to affect 168 out of every 10,000 children. Early detection of ASD is crucial as it allows for the implementation of early intervention strategies. These initial interventions have proven effective in mitigating deficits and can result in improved long-term outcomes for children. In the US, autism spectrum disorder affects approximately 3.63% of boys aged 3-17, compared to about 1.25% of girls [1] [2] [3].

Autism is a chronic illness that affects the patient and causes stress in the family. People with autism spectrum disorder have a high level of unemployment. Maintaining and obtaining employment is exceedingly challenging for young individuals with ASD, and some people with ASD have been qualified in paid job skills [4]. Medical imaging encompasses a variety of technologies that offer visible depictions of the inside of the human body to help radiotherapists and doctors discover, identify, and treat diseases more quickly and effectively. In recent years, deep learning has quickly become the technique of choice for clinical image interpretation [5].

Our brain is a complicated network comprising several functionally connected but spatially dispersed brain areas. In the past few decades, brain research has grown into a major community well-being concern. Research institutions and Scientists have devoted a significant amount of time and resources to improve understanding of the brain’s operating processes. Neuroimaging grew in popularity as research progressed [6]. Another class of autism spectrum disorder diagnosis methods that specialists prefer is neuroimaging approaches. Numerous investigations based on neuroimaging data, like functional and structural, for the diagnosis of ASD have been undertaken in recent years. Because it enables them to look at the structure and connections among different parts of the brain, structural brain imaging is a crucial technique for studying structural brain issues in autism spectrum disorder. Techniques utilizing magnetic resonance imaging are the main equipment used for structural neuroimaging. The analysis of brain activity and functional connections using functional neuroimaging can be helpful in the study of autism spectrum disorder. For fMRI, there are two techniques. For these, there are two terms: task-based functional magnetic resonance imaging and resting-state functional magnetic resonance imaging. ASD can be recognized using the rs-fMRI method [7]. Functional imaging techniques over structural approaches are more important for understanding ASD. Numerous medical disciplines are utilizing deep learning models, including structural and functional neuroimaging [9].

The blood oxygen level-dependent variations in magnetic resonance imaging indicate that occur when alterations in neural activity coincide with alteration in the state of mind are perceived by functional magnetic resonance imaging. The fMRI produces images of brain neuronal activity, which gives a unique and important piece of information for medical neurosciences, primary, and translational research. Screening and diagnosis approaches can be used to characterize autism spectrum disorder traits from a very early age, and similar techniques can also be used to determine the intensity of ASD. A variety of methods are utilized to check for autism spectrum disorder risk and severity, differentiate it from other neurodevelopmental disorders, plan therapy, and conduct research. There are no currently accepted treatments or medications for autism spectrum disorder [9] [10].

For many applications for medical image analysis, like segmentation, image classification, and registration, the preferred machine learning technique is quickly moving toward deep learning methods. Deep neural networks can learn directly from raw images and have a tremendous learning capability. As a result, learning meaningful models usually necessitates extensive training on big datasets and the application of appropriate model validation approaches. The huge data requirement makes interpreting many medical imaging datasets difficult. Clinical research uses deep learning algorithms to make disease diagnoses [10]. One of the most well-liked deep learning methods is the convolutional neural network. It was used to categorize a variety of domains, including heartbeats, the categorization of brain areas in MRIs, face detection, the recognition of race bib numbers, and license plate recognition [11]. Deep learning models are utilized in this study. The model we proposed for this study is VGG-20. Our study aims to distinguish children with ASD and children without ASD using a deep learning model [10] [12] [13].

## Literature Review

[5] proposed diagnoses of autism spectrum disorder by applying Deep graph convolutional neural networks (GCN). The structure was created on sixteen layers of graph convolutional neural networks. They joined scale data with neuroimaging with the use of autism brain imaging data exchange. They utilize a Deep graph convolutional neural network algorithm to use nodes with supervised learning designed for constructed graphs. For the training of this model, they used the drop edge strategy. While using Deep graph convolutional neural networks, their result displays 73% accuracy. [14] proposed a deep learning approach for using neuroimages to identify individuals with ASD. They used fMRI for the diagnosis of ASD. In their research, they utilized the Autism Brain Imaging Data Exchange (ABIDE). In their research, they used two models, ResNet-50 and VGG-16. The unprocessed data that were taken from ABIDE-I and ABIDE-II were three-dimensional; they converted these images into two-dimensional by using MRIcron software. A graphical user interface was also developed for the system. The accuracy acquired by these two models ResNet 50 and VGG 16, is 87% and 63% respectively. [15] presented deep learning models for the identification of ASD. In this paper, they create deep learning approaches for the classification of ASD. In their study, they used the ABIDE-1 dataset. Initially, they create brain systems from functional magnetic resonance imaging and describe the unprocessed features, like brain links. Then they utilize an autoencoder to learn unprocessed attributes. Then they train deep neural networks to achieve features. They apply an autoencoder and deep neural networks to achieve better accuracy. This model has an accuracy rate of 79%. [16] presented a deep learning model for the classification of ASD. They develop a model ASD-SAENet using fMRI data. They develop sparse autoencoder outcomes for obtaining features. Pearson’s correlation was used for feature selection. They develop an autoencoder for the rebuilding of features. They tested complete data and applied 10-fold layer cross-validation and obtained 70% accuracy. [16] presented an analysis and diagnosis of ASD with the use of Machine Learning. The dataset was gathered from the UCL ML repository. In the proposed technique, the Dataset was first normalized and cleaned to remove null values. After that, pre-processing was done on the data to split it into training and testing sets. SVM, Naïve Bayes, logical Regression, and Convolutional Neural Network (CNN) were applied to pre-processing data. Results showed that the proposed model detects and analyses ASD with an accuracy of 95%. [13] and [16] identified ASD using deep learning and the ABIDE dataset. UCI ML repository was used to gather the dataset. After that, encoding, normalization, variable reduction, and handling of missing data were done. A deep neural network model was applied to the data, and performance was compared with a support vector machine. The accuracy achieved by deep neural networks was higher than compared to SVM. Results showed that ASD can be diagnosed accurately by using Deep Neural Network. [17] presented deep neural networks for the diagnosis of ASD. Their approach consists of two prime aspects. Firstly, they built a deep neural network model with the use of TensorFlow. They used the ABIDE dataset. Second, using the same dataset and the same study designs, they used four distinct hidden-layer deep neural network models, each of which had been preprocessed using four different methods. They used four different arrangements of a multilayer perceptron. Their presented method has a 75.27 percent accuracy rate. [14] [18] presented a strategy for the automatic detection of ASD that combines a multi-atlas graph convolutional network and ensemble learning. To begin, they extract several feature representations from each subject’s fMRI data based on the functional connectivity of various brain atlases. This technique constructs several graphs based on the association among pairs of subjects, after which it applies the graph convolution process to these graphs. In this research, they make six graphs. Their presented method has a 75.86 percent accuracy rate. [19] provided CNN-based automatic diagnosis of ASD. They detected ASD with the use of fMRI. They employed a CNN architecture with one convolutional layer and tightly connected layers interleaved inside max-pooling. The output node is associated with a dense layer, which is then utilized to classify the data. The model was trained for 300 epochs with a batch size of 32, and the learning rate was set at 0.005. They acquired 70.22% accuracy, which is higher than the rest of the work. [20] [21] presented automated diagnosis of autism spectrum disorder were used to extract characteristics. In this research paper, they used a transfer learning approach. They utilized the VGG16 model for ASD detection for this purpose. The structural MRI images from the ABIDE dataset were used in this research. The VGG16 model’s last completely connected dense layer was replaced and retrained using extracted images from the ABIDE dataset. The researchers used the activation function softmax and optimizer ADAM to train the last dense layer with images with a 0.01 learning rate. The proposed approach had a 66 percent accuracy rate in detecting autism. [7] presented autism spectrum disorder detection using the outlier diagnosis method. A generative adversarial network was trained entirely with structural magnetic resonance imaging scans of healthy people to discover spatial patterns in the structural connections of the brain. The ABIDE-II dataset was used in this study. The goal of the generative adversarial network was to reassemble the following three consecutive segments from a stack of three adjoining portions as input. Their proposed work achieved an accuracy of 95.65%. [10] [20] [46] presented different methods of learning generalizable RNNs as fewer task-functional magnetic resonance imaging datasets. They used the data augmentation method. To create predictions of an entire-brain parcellation of functional magnetic resonance imaging data, they first use an RNN with extensive temporary memory. [45] [47] presented a deep learning methodology joint with the F-score attribute collection technique aimed at autism spectrum disorder identification. To extract a bottom characteristics representation that relates to the network’s bottleneck layer, an autoencoder is used. The prepared strategy was compared against two standard techniques: Stack autoencoder and ASD-DiagNet. Their suggested approach has the potential to achieve accuracy levels of 74%. [39]presented via 3D CNN and the temporal statistics of rs-fMRI data to classify autism spectrum disorder. They identified patients from the ABIDE I and ABIDE II datasets after pre-processing. They used a 3D convolutional neural network architecture in this research. To compare their 3D convolutional neural network models, they employed the linear SVM as a baseline. The accuracy of this method was 68%. [28] proposed a technique for comparing autistic patients to control participants utilizing combining sMRI and fMRI. They used the ABIDE-I dataset, and 817 samples were taken from the ABIDE-I dataset. After the dimension reduction phase, the first technique includes linking the attribute vectors, after then executing the autoencoder training. The second approach entails the use of independent classification processes, followed by a mechanism for selecting the final classifications. A multimodal approach using a group of classifiers for both functional and structural data classification led to the greatest classification accuracy of 5.06%. [4] proposed an artificial neural network model for the analysis of ASD. They provide a deep learning approach in this research paper to address the three issues with current techniques. First, by incorporating a convolutional layer that recognizes the combination of areas, they may deduce the parts of the brain connected for the classification of ASD. Secondly, they may discover the hidden layer of circulatory characteristics that alter over time by merging the temporal convolution layer with a recurrent neural network. Lastly, by including the details of the multiple sites, modeling the differences between the sites was another option. In their research, they used the ABIDE-I dataset. The existing model achieved an accuracy of 54% while their proposed model achieved ed accuracy of 74%. [16] presented based on brain structure, their proposed research intends to diagnose autism spectrum disorder from a big dataset by using CNN. Utilizing the feature extraction method, it determines the region of interest. In this research, the authors used the ABIDE-I dataset, which contains 539 autistic patients and 573 typical controls. A convolutional neural network analyzes the image attributes and picks up new information after a set number of training images. It uses matrix multiplication to apply filters on the input image’s pixel block. Each cell in the matrix has its Pearson correlation coefficient tested, and the results vary from -1 to 1. The learning rate was fixed at 0.005; likewise, the model’s training utilized 300 epochs and 32 batch sizes. Their model achieved an accuracy of 95%. [16] [17] proposed the Multi-site Clustering and Nested Feature Extraction (MC-NFE) technique for fMRI-based ASD detection. To simulate cross-site variations in every group, they train latent representations and execute subject grouping throughout every section using a multi-view feature model of continuous restoration. The ABIDE-I and ABIDE-II dataset was used in this study. In this study, the authors suggest an MC-NFE method for functional magnetic resonance imaging-based autism spectrum disorder identification that can handle high-dimensional fMRI data and reduce multi-variation. Their model achieved an accuracy of 68%.

## Background

### Magnetic Resonance Imaging (MRI)

MRI is a significant method for scanning the brain that provides high-resolution data on the structure, physiology, and working of the brain [34]. The brain imaging technique that accurately identifies the size and shape of the brain is magnetic resonance imaging [35]. The usage of MRI data to discover ASD indicators by revealing small changes in brain patterns and networks. Various researchers have found, by using MRI scans, that specific areas of the brain are related to ASD. According to the type of scanning method utilized, MRI scans are further classified into structural magnetic resonance imaging (s-MRI) and functional magnetic resonance imaging (fMRI) [31] [35].

**Table 1.** Details of Existing Literature.

| References | Classifier | Accuracy | Modality | Dataset |
| --- | --- | --- | --- | --- |
| [7] | DeepGCN | 73.7% | fMRI | ABIDE I<br>871 samples |
| [9] | VGG-16 and ResNet-50 | 63.4% and 87.0% | fMRI | ABIDE I, ABIDE II |
| [44] | autoencoder | 79.2% | fMRI | ABIDE-I<br>871 samples |
| [2] | ASD-SAENet | 70.8% | fMRI | ABIDE I 1035 samples |
| [11] | Convolutional<br>neural network model | 99.53%, 98.30%,<br>96.88% achieved<br>accuracy in three<br>datasets | Age, gender,<br>patient suffer from<br>jaundice, screening<br>test, screening score | ASD screening<br>in data for adults, children,<br>and adolescents |
| [12] | Deep neural network | 70% | rs-fMRI | ABIDE |
| [7] | Deep neural network | 75.27% | rs-fMRI | ABIDE 1035 samples |
| [13] | multi-atlas graph convolutional<br>network method (MAGCN) | 75.86% | fMRI | ABIDE<br>949 samples |
| [14] | Convolutional<br>neural network model | 70.22% | fMRI | ABIDE |
| [14] | VGG16 model | 55% – 65% | s-MRI | ABIDE |
| [16] | Generative Adversarial<br>Network (GAN) | 95.65% | T1w<br>s-MRI | ABIDE-II<br>38 samples |
| [8] | RNN with long<br>short-term memory | 52.5% | fMRI | Task-fMRI<br>40 samples |
| [17] | The deep learning approach<br>joined with the F-score feature<br>selection method | 70.9% | fMRI | ABIDE |
| [18] | Three-dimensional<br>convolutional neural network | 66% | rs-fMRI | ABIDE<br>1028 samples |
| [19] | Autoencoder and multi-layer<br>perceptron | 85.06% | fMRI,<br>s-MRI | ABIDE-I<br>817 samples |
| [20] | Artificial neural network | 74% | rs-fMRI | ABIDE-I<br>575 samples |
| [21] | Convolutional neural network | 95% | rs-fMRI | ABIDE-I |
| [13] | multi-site clustering<br>and nested feature extraction<br>(MC-NFE) | 68% | fMRI | ABIDE-I, ABIDE-II<br>609 samples |

#### Structural magnetic resonance imaging

To evaluate the neurology or anatomy of the brain, structural MRI scans are used. Additionally, structural MRI scans are used to quantify the size of the brain, including the volume of its subregions’ grey matter, and white matter [31]. Structural MRI may be used to acquire anatomical characteristics of the brain, during brain MRI analysis [34].

#### Functional magnetic resonance imaging

To see the active brain parts linked to brain function, functional MRI (fMRI) scans are used. By detecting variations in blood flow across several cognitive areas, f-MRI calculates synchronized activity in the brain [31]. fMRI measures the Blood Oxygen Level Dependent Signals’ temporal connections with various parts of the brain when the individual is at rest from cognitive activity. To analyze connectivity and to distinguish ASD patients from neurotypical people, the pairwise correlation coefficients of the Blood Oxygen Level Dependent sign obtained from the numerous parts of the brain were directly utilized [35].

#### Resting-state Functional magnetic resonance imaging

Rs-fMRI has emerged as a vital method for studying neurology. Investigating the variations in activities of the brain between people with autism spectrum disorder and healthy control subjects makes it simpler to identify the underlying causes of ASD, resulting in a more accurate diagnosis and path of treatments. The brain’s regions are represented by the graph’s nodes, and its edges show how those nodes are connected [42]. According to conventional fMRI procedures, each element of the brain’s cognitive system must be studied separately. A relatively new technique, resting-state functional connectivity, may have the benefit of overcoming constraints in traditional fMRI techniques [44].

### Deep learning

The development of high-performance computing resources led to the popularity of deep learning approaches that use these networks. Deep learning is implemented nowadays in several applications, such as image identification, vision, and language processing. The data is passed through numerous layers of the deep learning algorithm; each layer can gradually extract the features and pass the data to the next layer [21]. As the big data period has developed, deep learning has advanced to have a more complex network structure and more powerful feature learning and characteristic representation abilities than traditional machine learning algorithms. Deep learning methods are used by the most effective classifiers that successfully categorize images [24]. Medical imaging is a field where deep learning technology is extremely useful. One of these fields that has been greatly impacted by this quick advancement is medical image processing, particularly in image identification and classification [20]. The following are the deep learning models

#### Convolutional Neural Network

Every layer in a convolutional neural network uses a discrete feature to convert one volume of activation functions into another. Pooling layers are placed between the convolutional layers, which ultimately results in fully linked layers [48]. CNN has emerged as a significant tool for analyzing medical images. Using techniques involving image classification, localization, segmentation, detection, and more, CNNs make a significant advancement in the domain of analyzing medical images. The fundamental components of a Convolutional Neural Network include convolutional, pooling, and fully connected layers, and activation functions [30].

#### Recurrent Neural Network

RNNs manage the data in a different way than CNNs by using time-series knowledge and processing it in situations with consecutive input and disrupted response. It analyzes sequential input, picks up features from that sequence, and outputs another sequence. It belongs to the ANN family and consists of input, output, and hidden nodes [8]. For processing sequence data, RNNs have also been constructed [3]. An extension of an RNN may extract the features and long-term connections from consecutive and time information [48].

#### Autoencoder

An autoencoder is an unsupervised algorithm for learning that compresses input into a latent-space representation using the backpropagation technique with predicted values that are identical to the inputs. It consists of two sections: The network’s encoder section reduces the input into a representation of latent space. decoder section recreates the input from the latent space model [12]. Several autoencoders are layered together to make a deep system. Additionally, several types of autoencoders, sparse autoencoders, denoising autoencoders, and variational autoencoders [48].

### Visual Geometry Group (VGG)

In [31], K. Simonyan and A. Zisserman proposed the VGG. It is one of the well-known deep-learning architectures. In 2014, the VGG, a common form of convolutional neural network, was first proposed. To extract image characteristics, it has been mostly used. The increase in the depth of neural networks can enhance the performance of the network [43]. VGG is a common deep CNN design with multiple layers. With VGG-16 or VGG-19 having 16 or 19 convolutional layers, respectively, the word “deep” refers to the number of layers.

#### Visual Geometry Group-16 (VGG-16)

The ImageNet database was used to train the VGG-16. The VGG-16 has undergone significant training, which results in good accuracy even with small image data sets [38]. It takes low-level input images and runs them through a series of computing units to get the results it needs for higher-layer categorization. The convolutional layers extract features from the input data. It is a large model with about 138 million parameters. Five blocks comprised the 13 convolutional layers. The first two phases each contain two convolutional layers. The final three blocks each have three convolutional layers [6].

#### Visual Geometry Group-19 (VGG-19)

VGG-19 is a variant of the VGG structures that has 19 highly connected layers. VGG19 has 16 convolutional and 3 fully connected layers. The model’s convolutional layers are fully and firmly linked, which enhances the extraction of features and enables the use of Maxpooling before identification using the activation function [15]. The complete convolution section is divided into five sub-regions by five max-pooling layers in a sequence. The first and second subregions are made up of two convolution layers, each with a depth dimension of 64 and 128. The remaining three convolution layers have depth sizes of 256, 512, and 512. The output element is positioned between two hidden layers. To minimize overfitting in its implementation, L2 regularization is used in this model after every completely connected layer [23].

## Methodology

To classify autism deep learning model, VGG20 has been proposed. VGG 20 consists of 20 layers. The dataset Autism Brain Imaging Data Exchange I (ABIDE-I) was used in this research. The dataset was downloaded via their website, Autism Brain Imaging Data Exchange, in NIFTI format. The main steps of the methodology consist of converting NII images to PNG format, pre-processing, converting images to the array, training the model, and displaying results. In a preprocessing step, normalization is applied. The following figure shows the proposed methodology.

### Dataset

ABIDE-I dataset is selected for the training of the proposed model.

#### ABIDE-I Dataset

The most prominent data-driven method for autism identification and biomarker research is the autism brain imaging data exchange (ABIDE), a joint effort that incorporates brain imaging and phenotype data collected from 1,112 subjects. The ABIDE I includes 1,112 participants from 17 sites, including 539 ASD patients and 573 others. In our research, we used the ABIDE-I dataset. The ABIDE I dataset is made up of structural and resting-state functional magnetic resonance imaging data in addition to phenotypic information collected from 17 international imaging locations [32].

**Table 2.** Demographics Information of ABIDE Dataset.

| Site ID | ASD | HC | Male | Female | Age at scan |
| --- | --- | --- | --- | --- | --- |
| CALTECH | 19 | 19 | 30 | 8 | 17.0-56.2 |
| CMU | 14 | 13 | 21 | 6 | 19-40 |
| LEUVEN | 29 | 35 | 56 | 8 | 12.1-32 |
| MAX_MUN | 24 | 33 | 50 | 7 | 7-58 |
| OHSU | 13 | 15 | 28 | 0 | 8.0-15.2 |
| OLIN | 20 | 16 | 31 | 5 | 10-24 |
| PITT | 30 | 27 | 49 | 8 | 9.3-35.2 |
| SBL | 15 | 15 | 30 | 0 | 20-64 |
| SDSU | 14 | 22 | 29 | 7 | 8.7-17.2 |
| STANFORD | 20 | 20 | 32 | 8 | 1.5-12.9 |
| TRINITY | 24 | 25 | 49 | 0 | 12.0-25.9 |
| UCLA | 13 | 14 | 25 | 2 | 8.4-17.9 |
| YALE | 28 | 28 | 40 | 16 | 7.0-17.8 |

### Steps for Converting NII Images into PNG

Initially, the dataset is present in NII format so we need to convert images into PNG format. Here are some steps for the conversion of images.

#### Extraction of NIFTI files

The dataset that we have obtained (i.e., ABIDE-I); is currently in Raw Form. These raw format files are NIfTI imaging format, which is a way to look at objects as 3D images in art and with other neural imaging software. NIfTI stands for Neuroimaging Information Technology Initiative. It’s a standard representation of images, and it’s the most commonly used type of analytic file. These files have been developed to facilitate cross-platform and cross-software interpretability. These images are 3-dimensional arrays. The file representation for the NIfTI is a 3D image of the entire brain, or a 3-dimensional array of numbers. The header for the NIfTI doesn’t contain patient information, but it does have imaging metadata such as the pixel dimensions.

#### Creating Folder

For NIfTIs, since they only have one image for each file, we can store these all together for one subject in a single folder. For this, we create a folder. The extracted data consists of two folders, which are autism and non-autism.

#### Extraction of Files

After extracting the files in the folder, there’s a package that’s created for working with NIfTI files called Oro.NIfTI. And this has a function called writeNIfTI and readNIfTI which has been used in many analyses. Each NII file moves into the corresponding folder.

#### Converting NII to PNG

As PC can’t be viewed directly NII images, there is a need to convert each image into (PNG/JPG format) using code. There is also some software available that converts NII images into PNG, like MRIcron. MRIcron is a 3D viewer and volume renderer for medical images. It is designed for NIfTI format images (popular with neuroimaging scientists) but can typically view images in many popular 3D raster formats including BioRad PIC, DICOM, NRRD, MGZ/MGH, AFNI BRIK, ITK MHA format images. The drawback of this software is that it can only convert 1 image at a time. So, it’s best to write some code that would convert the images in bulk. So, we made a code that helped us to convert NII to PNG images.

#### Images to Array

In Python, the numpy library reads the images in array form, so we need to convert the images into an array format. We used Python with the numpy library to convert 3D images into 2D images. When we give the path of images to the library, it automatically converts into an array. This array ranges from 0 to 255 values.

### Pre-Processing

All the changes made to the original data before feeding it to the deep learning algorithm are referred to as pre-processing. The errors from the rs-fMRI data are removed using the same preparation techniques as (Mostafa, S, Tang, L, et al., 2019), which prepares the data for the subsequent ROI extraction stages. Pre-processing comprises Cleaning, batch normalization, and noise removal. Cleaning and cropping the images was the goal of the data preparation. The fact that the data were gathered from online sources meant that they needed to be pre-processed before being utilized. Preprocessing is commonly implemented to transform data into the proper type, standardize the data in some way, or extract relevant information. 90% of the dataset for our research study was used for training, 5% for testing, and 5% for validation.

### Normalization

Normalization is a process in which data are transformed to be on the same scale. Normalization is the process of converting the numeric columns of a dataset to a standard scale without eliminating any data or affecting the value range. It removes any extraneous data. The dataset was rescaled with all image parameters from [0, 255] to [0, 1] using the normalization technique.

## Proposed Model

This study proposed the VGG20 model and used the ABIDE-I dataset for the detection of autism and non-autism. The Visual Geometry Group Network (VGG20), a 20-layer deep artificial neural network model, is referred to by the term VGG20. 16 convolutional layers and 4 fully connected layers make up the VGG20 model. Figure 5.2 shows the architecture of the VGG20 model.

**Fig 1.**
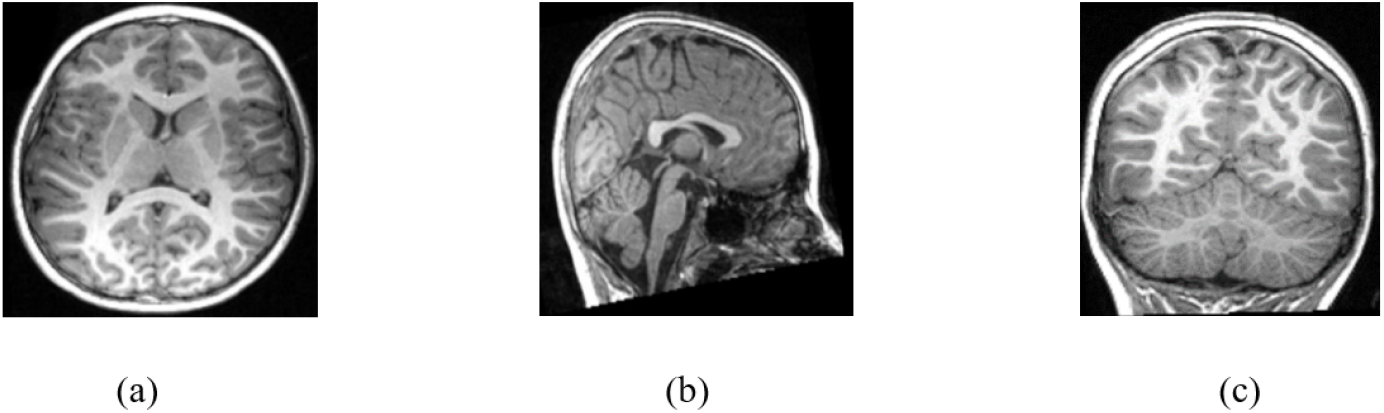
(a) Axial View (b) Sagittal View (c) Coronal View

**Fig 2.**
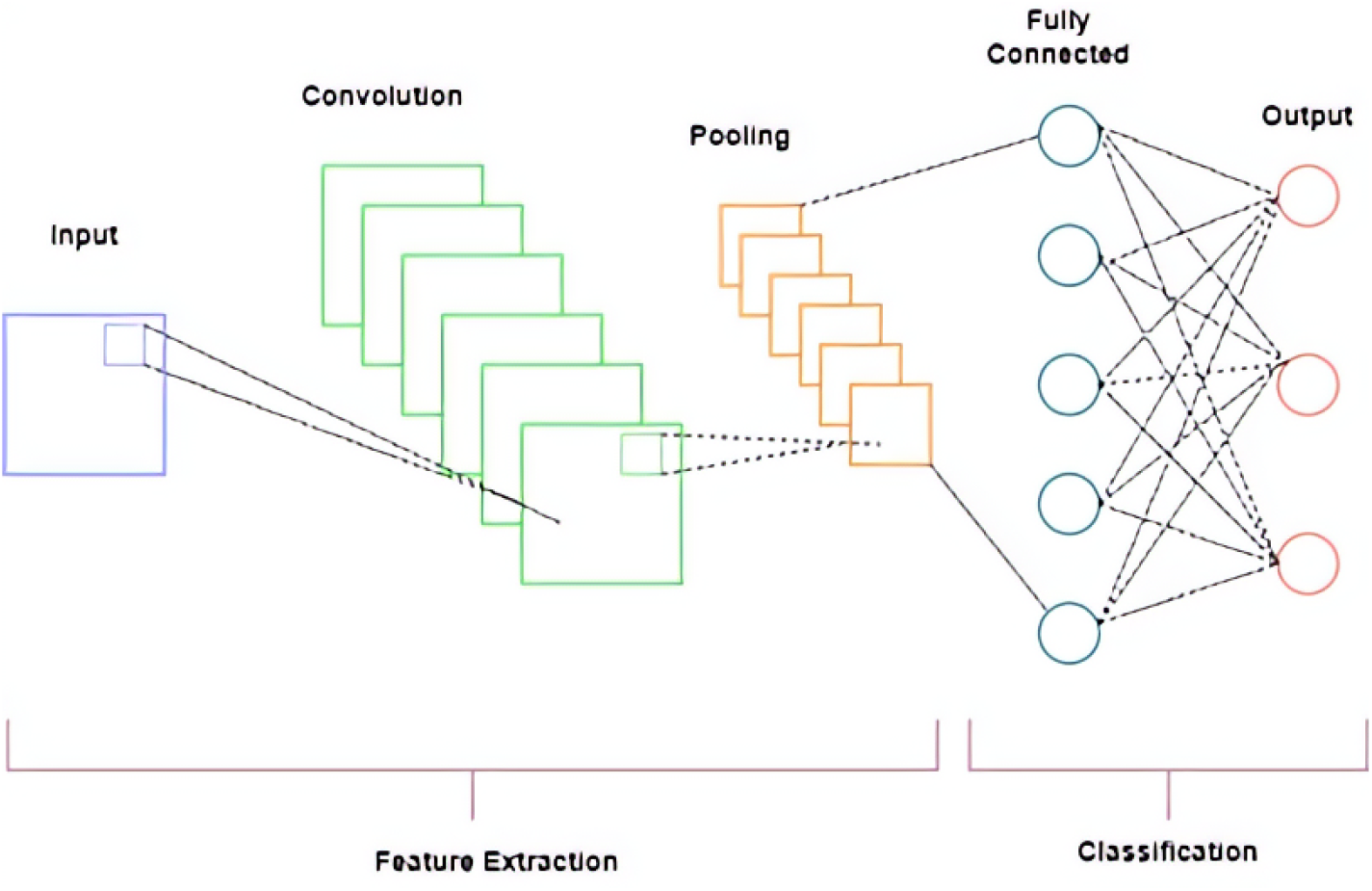
Architecture of CNN [30]

**Fig 3.**
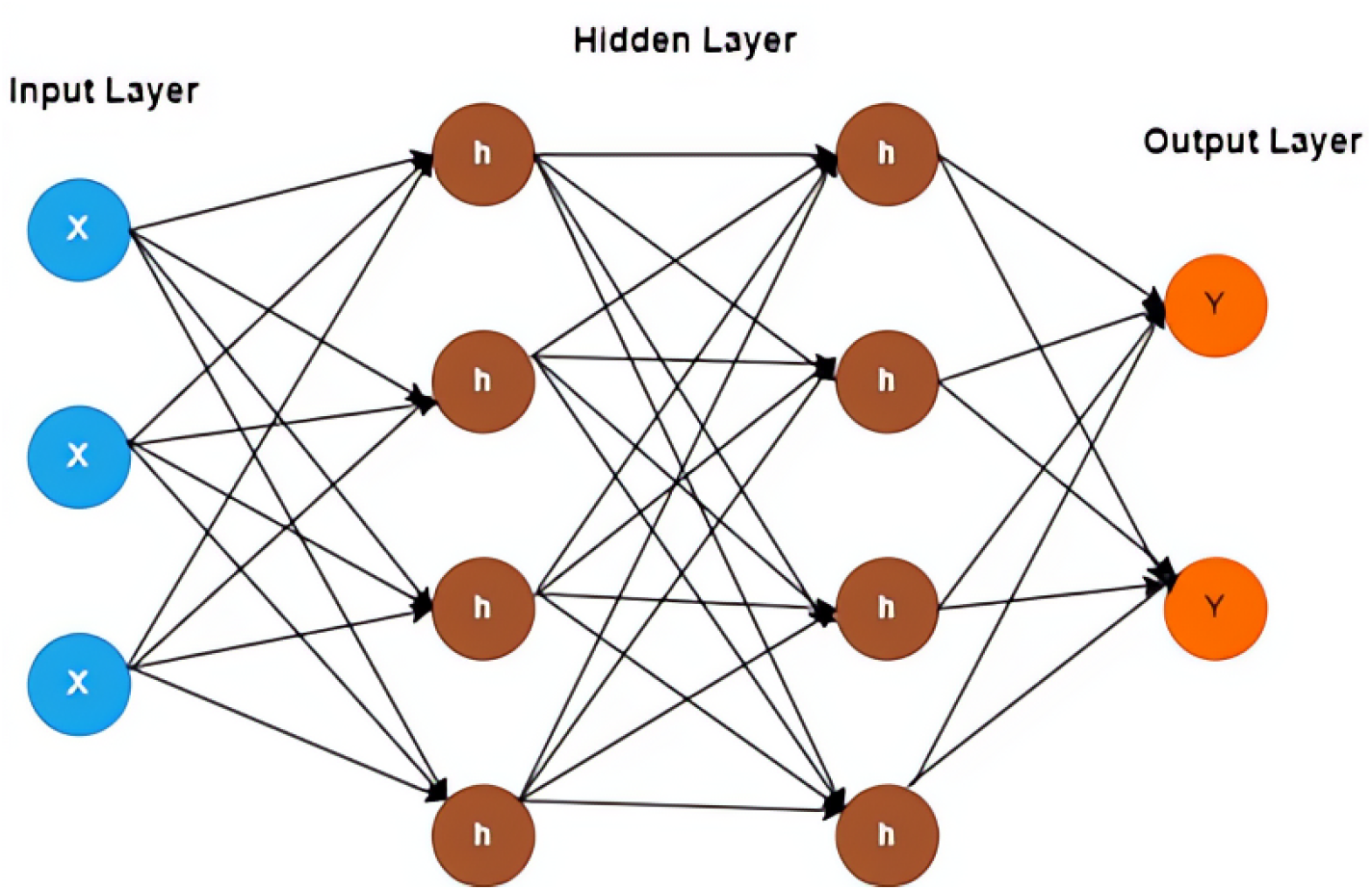
Architecture of RNN (Katte, T. 2018)

**Fig 4.**
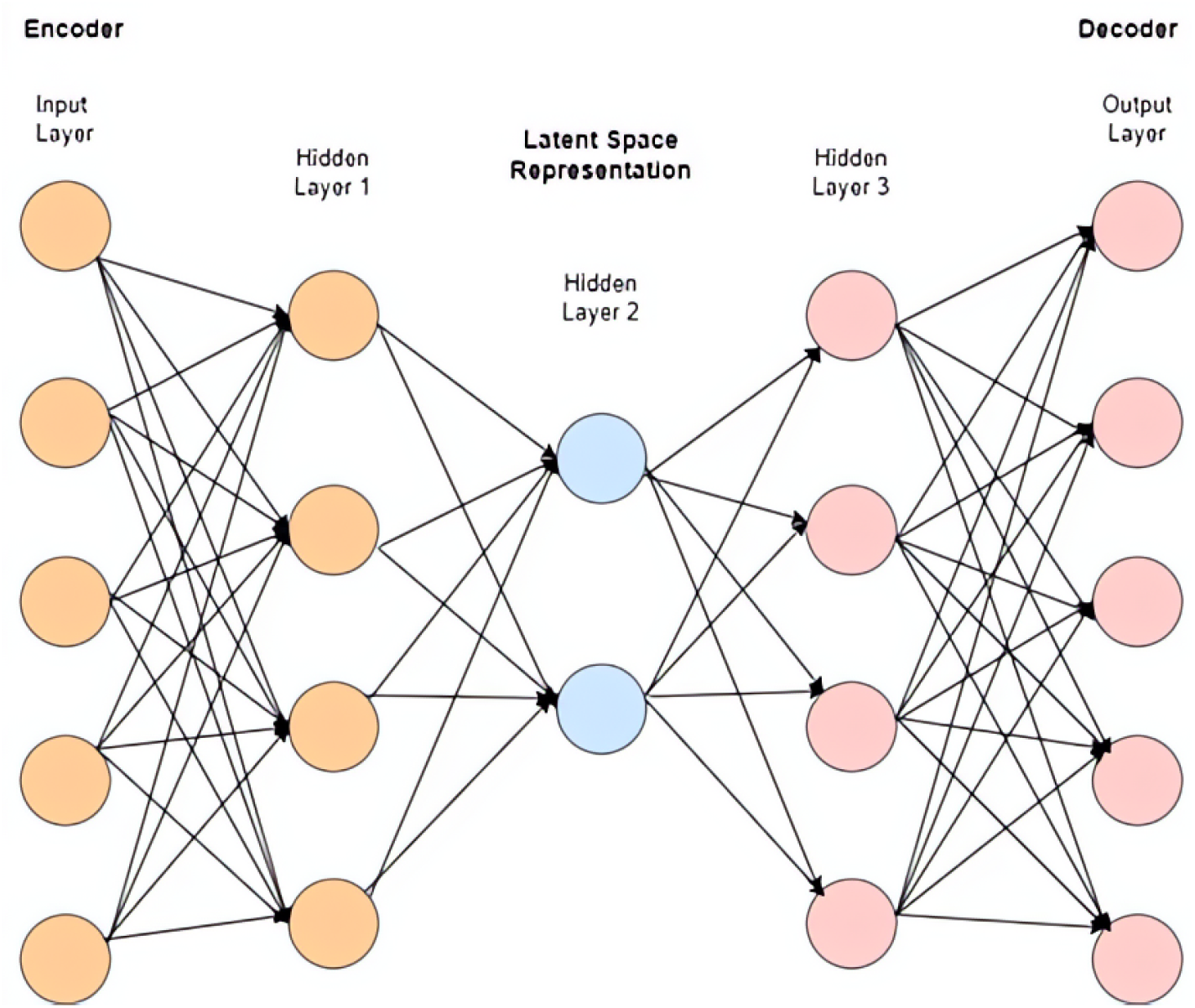
Architecture of Autoencoder [46]

**Fig 5.**
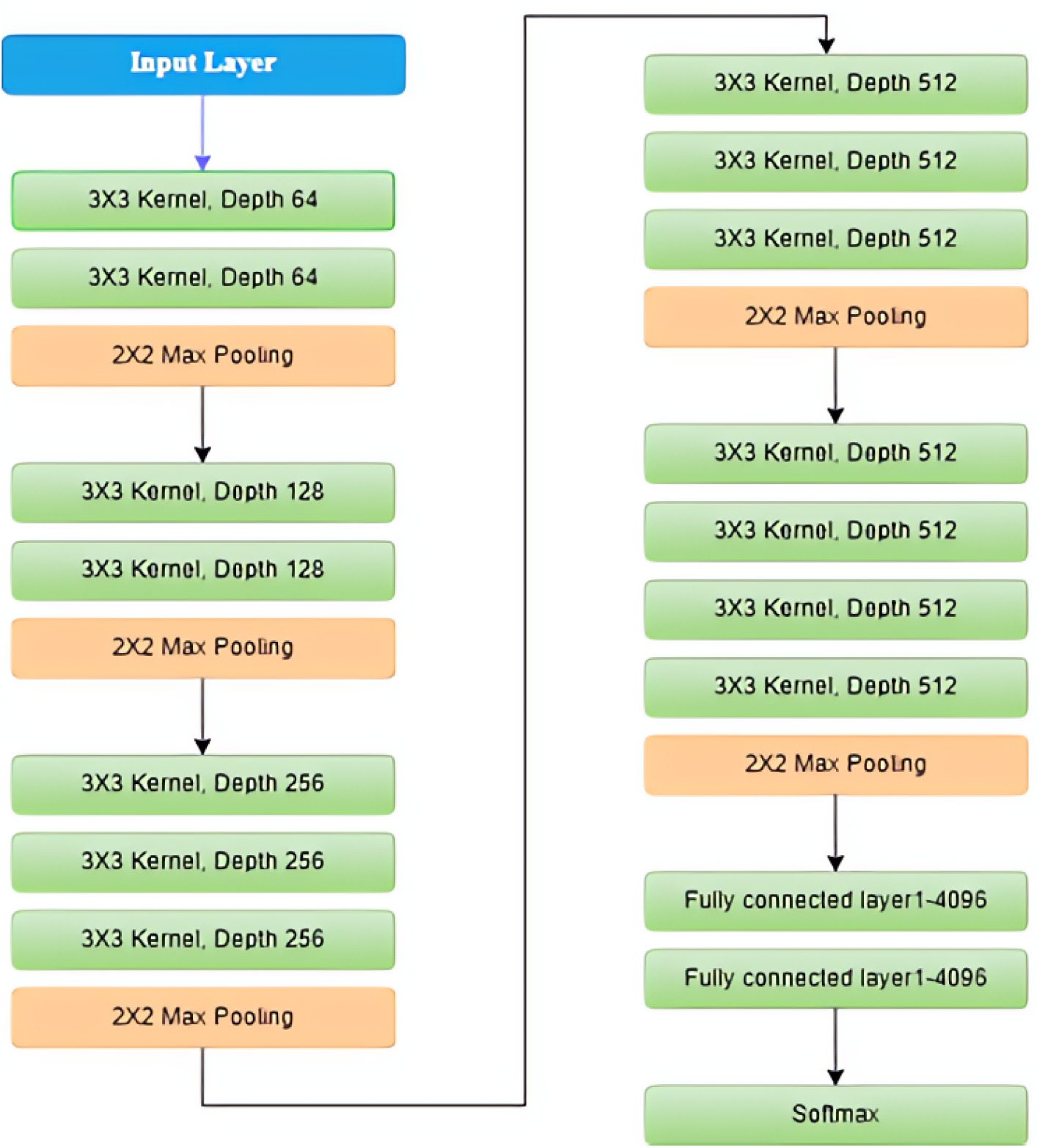
Block diagram of VGG-16 [37]

**Fig 6.**
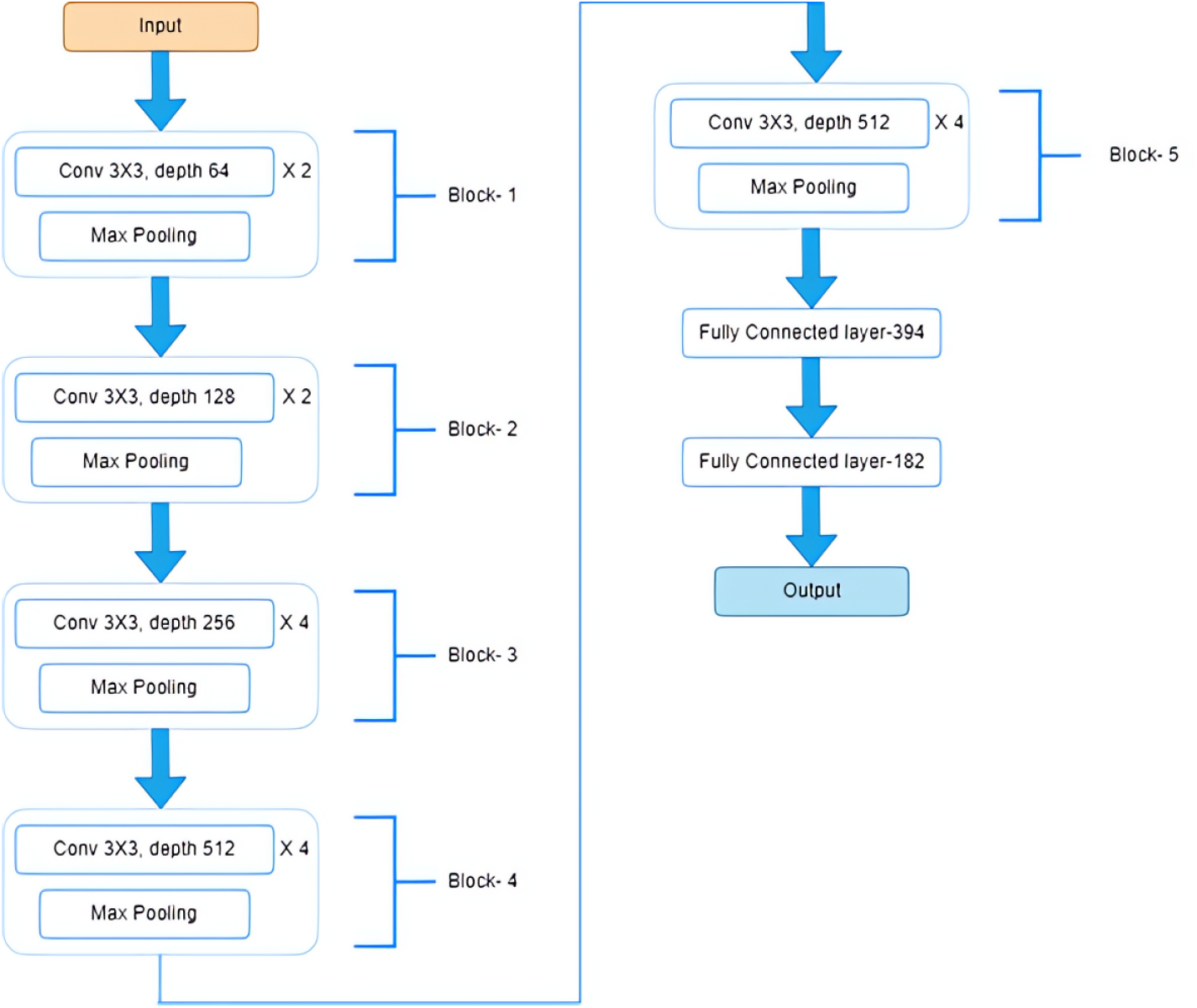
Block diagram of VGG-19

**Fig 7.**
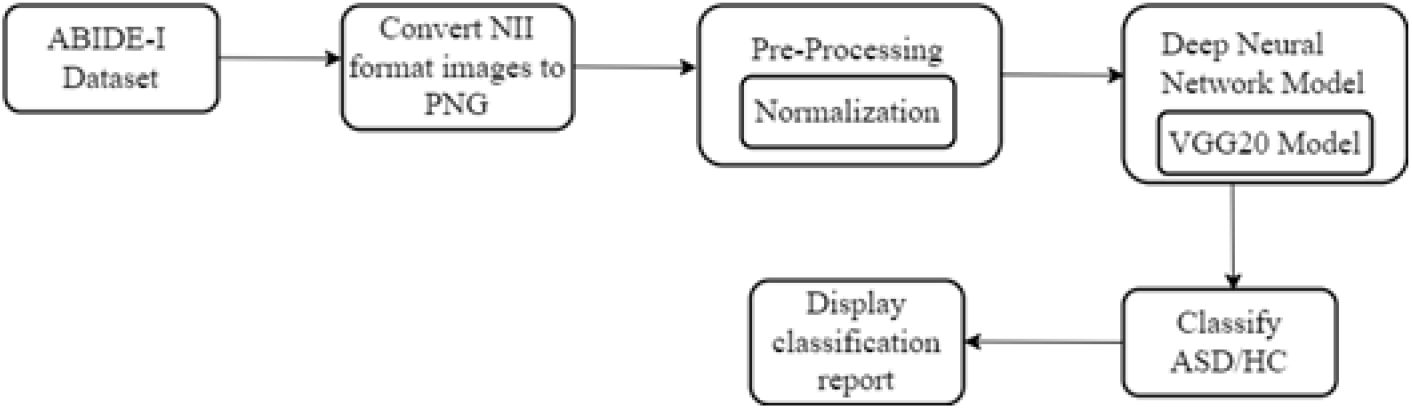
Proposed Methodology

**Fig 8.**
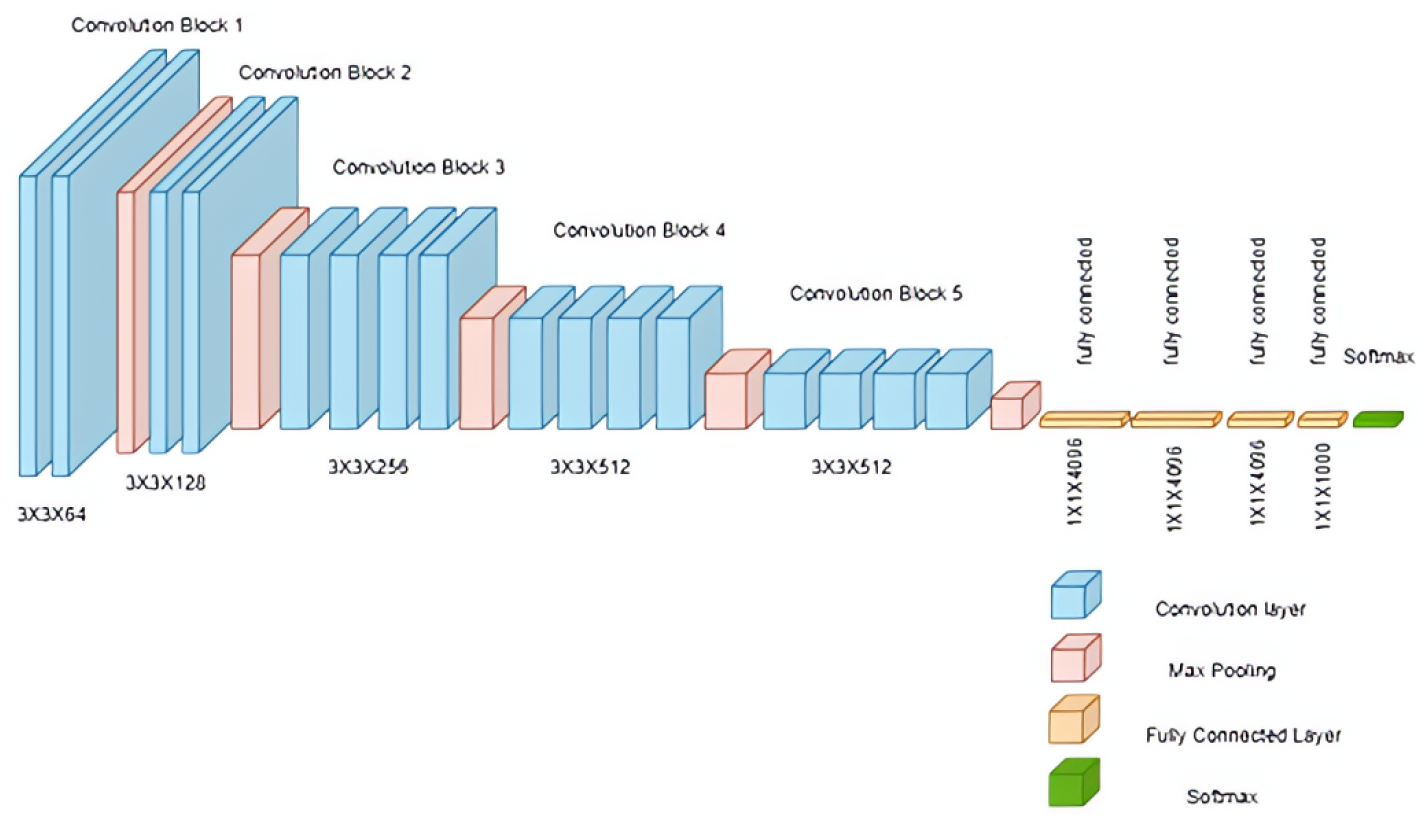
Architecture of VGG20 model

On the ImageNet dataset, the VGG20 algorithm which essentially draws on the convolution neural network approach, is often used and is useful for its simplicity because it has three convolutional layers connected to its upper side that rise with changes in gravity. In VGG20, max pooling layers were used as assigners to minimize the input volume size. Three FC layers with a total of 4,096 neurons and one layer with 1000 neurons were used in the VGG20 structure to connect each layer to the others The input layer’s purpose is to receive an image input with the dimensions 224, 224, 3. The central 224, 224, 3 patches in every image were selected by the model’s developers for the sake of keeping the image consistent in the input size for the ImageNet competition. VGG20 model contains 16 convolution layers. The convolutional layers of the VGG use a small receptive field, or 3X3, which is the lowest practical size that nevertheless releases both up and down as well as left and right. The convolution stride is set at 1 pixel to keep the spatial resolution after the convolution. All the hidden layers of the VGG network use ReLU. VGG rarely affects Local Response Normalization because of its longer training time and memory requirements. Additionally, it does not improve the model’s overall accuracy. The first three fully connected layers of the VGG20 each feature 4,096 channels, and layer four has 1,000 channels.

**Table 3.** Hyperparameter for Proposed Model.

| Hyperparameters | ABIDE-I Dataset |
| --- | --- |
| Batch Size | 32 |
| Number of Epoch | 20 |
| Learning Rate | 0.001 |
| Optimizer | Adam |
| Convolution Kernel Size | (3, 3) |
| Max Pooling Kernel Size | (2, 2) |

### Data Augmentation

Machine learning methods, particularly deep learning techniques, can be helpful if given enough training data. The model overfits and is not generalizable due to insufficient data [29]. Huge training sets are often not accessible, and gathering additional data might be expensive, similar to how medical imaging works. In these cases, a training set can be utilized with data augmentation techniques to create simulated data. To expand the amount of data or produce artificial data, data augmentation involves adding copies of previously existing data and making minor modifications to them [25]. For various applications, several data augmentation techniques have been proposed, including random translation, rotation, and cropping, and adding random noise to the features. In this study, the cropping augmentation method was used.

### Performance Evaluation

Evaluation measures are used to evaluate how well the statistical or deep learning model is performing. To assess the model’s quality, evaluation measures are applied. Before we put our model into production on untested data, we should be able to increase its overall predictive power by assessing its performance using several metrics. A few metrics used to evaluate the performance of the model include recall, precision, accuracy, and F1-score.

## Results

### Training and Testing Accuracy

In this section, the findings of the research work performed to classify autism spectrum disorder. One pre-trained deep learning model, the VGG20, was used in these studies to identify ASD. Based on the neuroimaging characteristics of the children, a deep learning model was developed and evaluated to identify the characteristics that distinguish autism and non-autism. Figure 9 and 10 depicts the accuracy and loss graph, with the x-axis showing the epochs while the y-axis shows accuracy. The dataset was split into training, validation, and testing with a ratio of 90:5:5. The training accuracy of the proposed model is 90%, validation accuracy is 69% and testing accuracy was 61%. The VGG20 model accuracy increased during training, going from 57% to over 90% after 6 epochs. Its validation loss was 0.60, while its training loss was 0.72.

**Fig 9.**
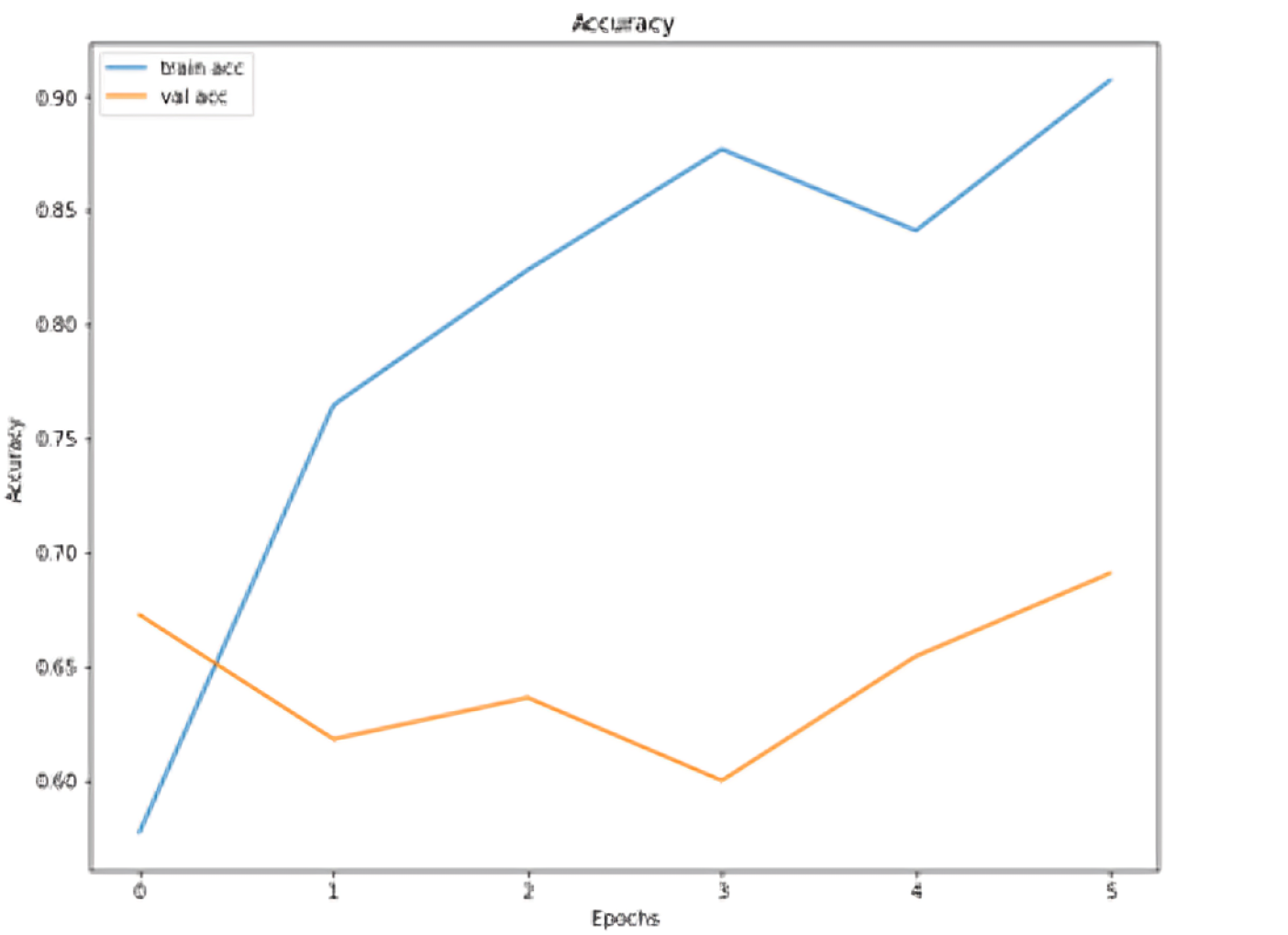
Model performance plot for the VGG20 model

**Fig 10.**
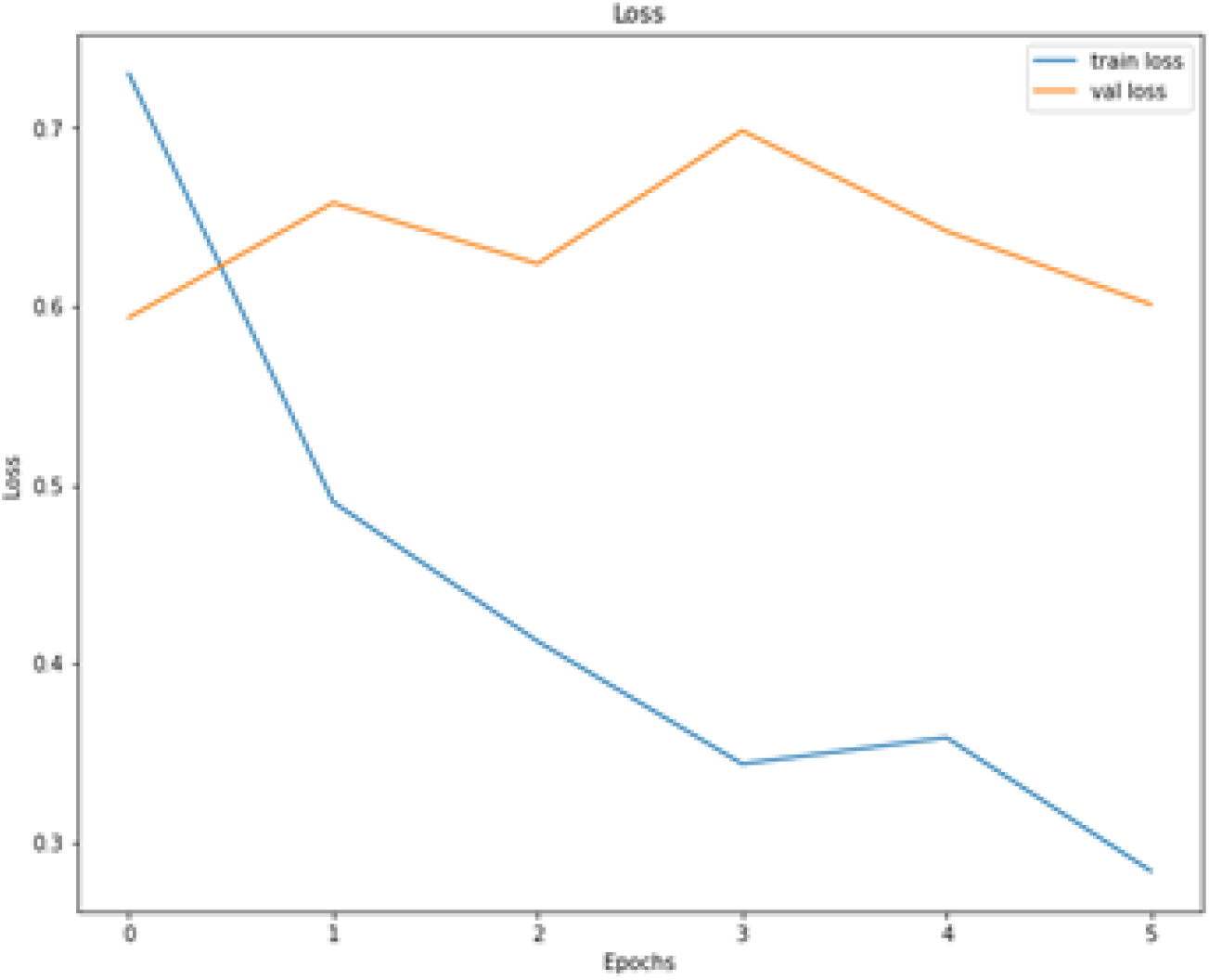
Model loss graph plot for the VGG20 model

**Fig 11.**
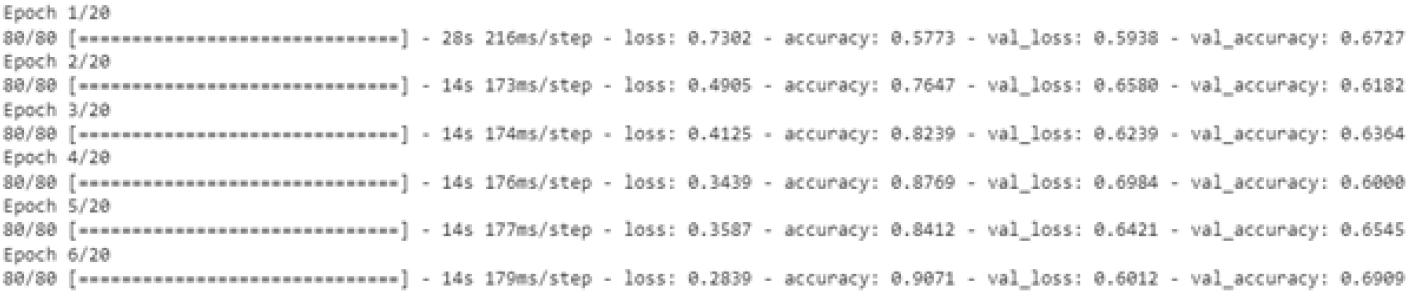
Number of Epoch for VGG20 model

### Training and Testing Accuracy

In this section, the findings of the research work performed to classify autism spectrum disorder are presented. One pre-trained deep learning model, the VGG20, was used in these studies to identify ASD. Based on the neuroimaging characteristics of the children, a deep learning model was developed and evaluated to identify the characteristics that distinguish autism and non-autism. Figures 9 and 10 depict the accuracy and loss graph, with the x-axis showing the epochs while the y-axis shows accuracy. The dataset was split into training, validation, and testing with a ratio of 90:5:5. The training accuracy of the proposed model is 90%, validation accuracy is 69% and testing accuracy was 61%. The VGG20 model accuracy increased during training, going from 57% to over 90% after 6 epochs. Its validation loss was 0.60, while its training loss was 0.72.

The pre-trained model had a 61% accuracy rate using the 32-batch size as shown in Figure 12.

**Fig 12.**
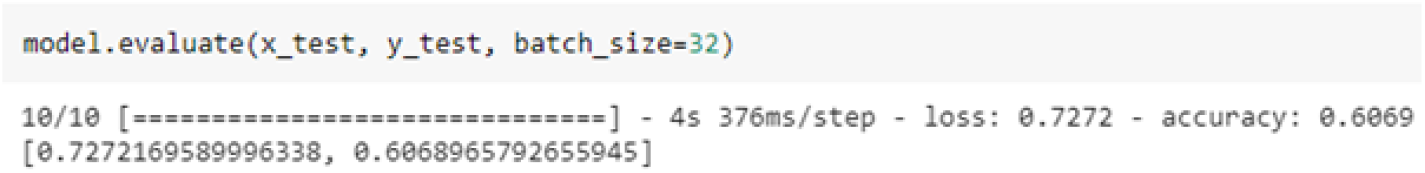
Result of Evaluation Metrics

Moreover, the evaluation matrices like Precision, f1 score, recall, and score achieved findings are displayed in Figure 13. The figure exhibits that Autism precision is 70% which is more than the non-Autism percentage, recall is 59%, which is less than the non-autism percentage, f1-score accuracy is 64%, and accuracy lies at 61%.

**Fig 13.**
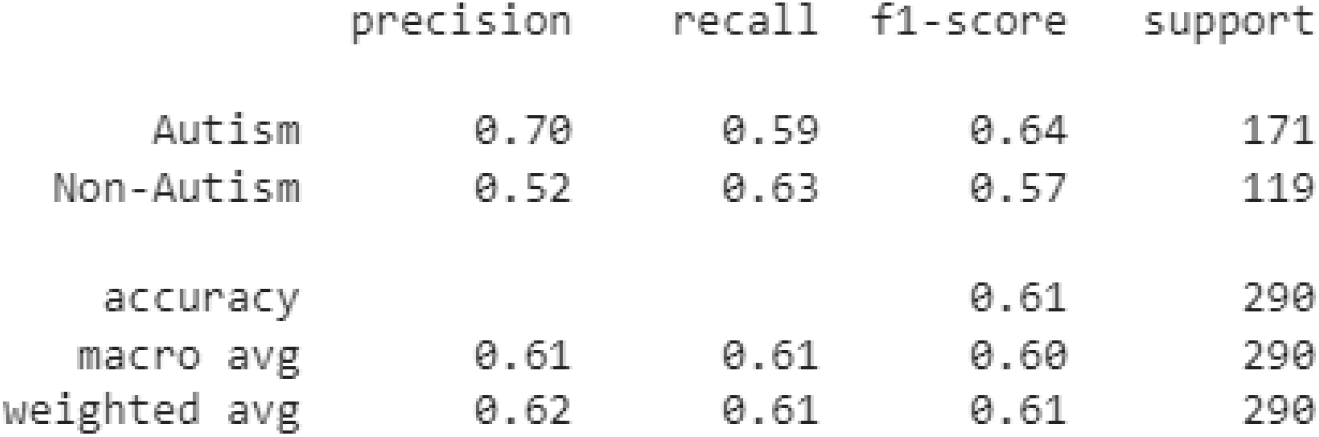
Accuracy result of the VGG20 model

The VGG20 deep learning model’s confusion metrics are displayed in Figure 14. The figure shows that the model predicted the true values of autism are 101 while the non-autism values are 75.

**Fig 14.**
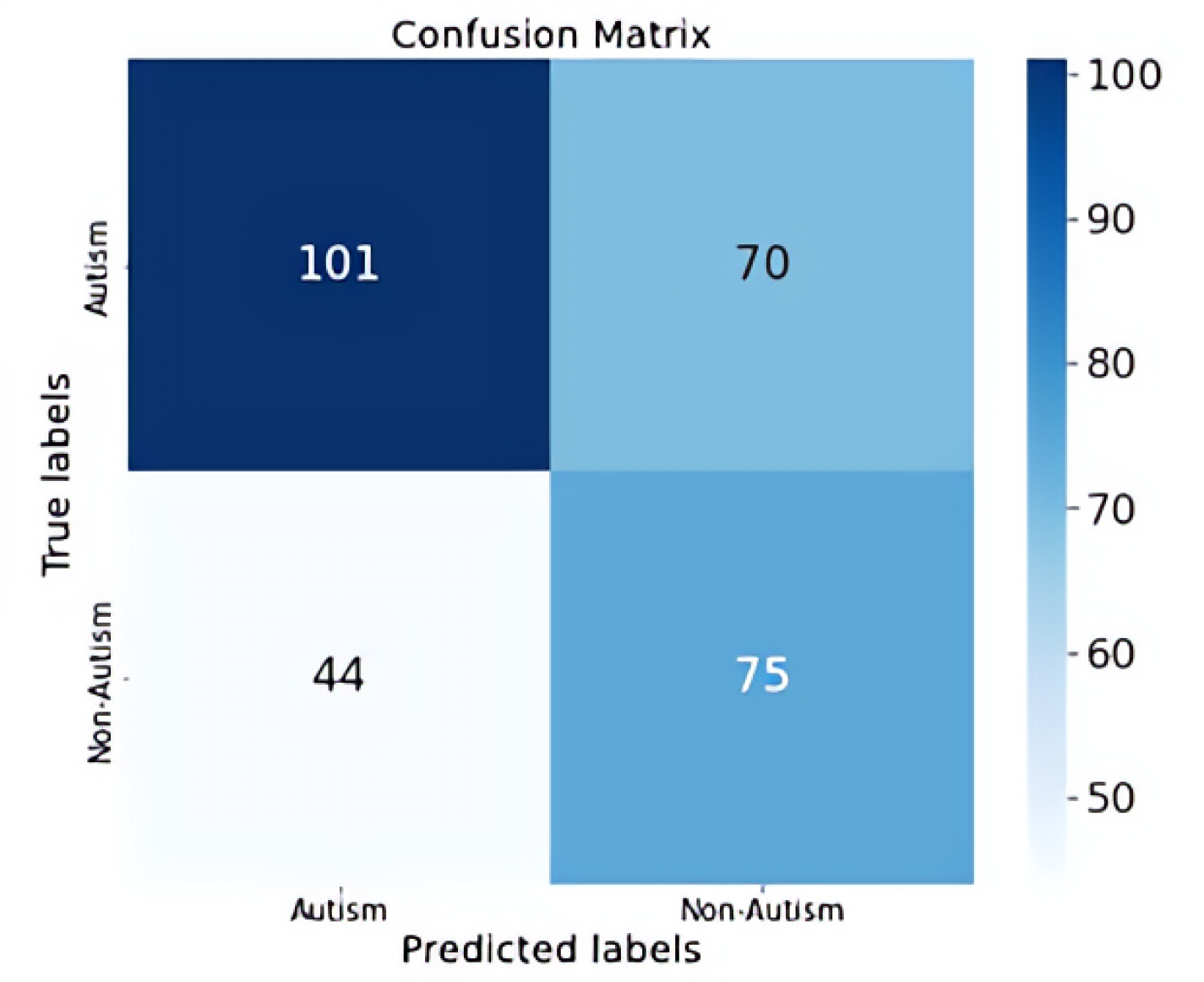
Confusion Matrix for VGG20 model

### Performance Comparison

presented an inspection Resnet V2 model. In this research, the author used the ABIDE-I dataset. From this dataset, they used 172 samples in their research. Their suggested model had a 57% accuracy rate. [42] presented the classification of ASD by using an RNN with a long short-term memory model. The researchers utilized a small task functional magnetic resonance imaging dataset. They took only 40 samples from this dataset. Their model has a 52% accuracy rate. [45] developed a deep autoencoder model to identify ASD. In this research paper, the authors used the ABIDE-I dataset. They took only 25 samples from this dataset. 54% accuracy was attained by the suggested model. [8] presented a machine learning model. In this research, the authors proposed a support vector machine model. On the ABIDE dataset, their model had a 52% accuracy rate. The authors took 341 samples for their research.

## Discussion

The term ASD is used to characterize a variety of repeated visual and auditory behavioral patterns and early-appearing social communication deficits that have genetic factors roots as well as other factors. Nowadays, autism is thought to exist on a spectrum with varying degrees of severity. Genetics and neurology have discovered significant risk factors, but there haven’t been many practical uses until now [19]. According to a survey, autism affects 1 in every 70 children globally. Autism spectrum disorder affects roughly 3.63% of boys aged 3 to 17 in the US, in comparison to about 1.25% of girls [1]. Various medical disciplines are utilizing deep learning algorithms, including structural and functional neuroimaging [1]. For many medical image analyses, applications/programs such as segmentation, deep learning approaches, image classification, and registration, are rapidly replacing existing machine learning methods. Deep neural networks can learn directly from raw images and have a tremendous learning capability [10]. Our study’s objective was to distinguish children with ASD and children without ASD using deep neural networks. The autism brain imaging data exchange (ABIDE 1) dataset was used in this study. The most popular information approach aimed at autism identification and biomarker research is the autism brain imaging data exchange (ABIDE), a joint project that incorporates brain imaging and phenotype data collected from 1,112 individuals. The author used 2725 images from the ABIDE-1 dataset for her research. The ABIDE I is made up of anatomical and resting-state fMRI data in addition to phenotypic information collected from 17 international imaging locations [32]. The dataset that the researcher had obtained (i.e., ABIDE-1), was in raw form, i.e., NIfTI. It’s a standard representation of images, and it’s the most commonly used type of analytic file. The file representation of the NIfTI is such that it has a 3D image of the entire brain or a 3-dimensional array of numbers. The author had to convert NIFTI images into a format that is easily readable for a personal computer, i.e., PNG format. That’s why the author had to first convert the entire dataset to the PNG format. Then the researcher had to apply normalization for preprocessing of images. The challenge was of rescaling all of the picture/image parameters from [0, 255] to [0, 1]. Furthermore, the author for the classification of the images, utilized deep learning models in her study. The model that the researcher proposed and implemented was VGG-20. The name VGG20 refers to the visual geometry group network, which contains 20 layers. Consequently, ABIDE 1 dataset images were not sufficient for training the model, so to counter this problem, the researcher applied the “cropping data augmentation technique” to expand the dataset. The optimizer used in this study’s model was Adam with a batch size of 32. Similarly, for the training of the model, the author used 20 epochs. In this study, recall, accuracy, F1 score, and precision were the four evaluation metrics used. The suggested model thus attained a 61% accuracy.

**Table 4.** Comparison of the Existing model with the Proposed model.

| Reference | Model | Accuracy | Dataset |
| --- | --- | --- | --- |
| [41] | Inspection Resnet V2 | 57% | ABIDE-I<br>172 samples |
| [42] | Long short-term memory (LSTM) | 52.2% | Small Task fMRI<br>40 samples |
| [45] | Deep autoencoder | 54% | ABIDE I<br>25 samples |
| [8] | Support Vector Machine | 52% | ABIDE<br>341 samples |
| Proposed Model | VGG20 | 61% | ABIDE-I |

## Conclusion and Future work

Autism is indeed a neurological condition characterized by severe as well as continuous behavior patterns that appear early in development, issues with social communication and interaction, and other signs, behavior, and interests. In this research, the difficulty of differentiating between individuals with ASD and healthy volunteers was addressed. In contrast to previous investigations by other authors, who used a range of machine-learning models, this study was conducted using a deep-learning model. The VGG20 model for the classification of autism spectrum disorder was also proposed and implemented by the researcher using the autism brain imaging data exchange 1 (ABIDE 1) dataset. This study might be useful for categorizing autistic and non-autistic patients. Our model had a 61% accuracy rate. Future recommendations include creating new deep-learning techniques and training them using various datasets, such as ABIDE 2. Future researchers can also include the varied viewpoints of the neuroimaging dataset.

## Data Availability

All data produced in the present work are contained in the manuscript

## Acknowledgments

This research is based on the study of neuroimaging-based diagnosis of Autism Spectrum Disorder. The authors gratefully acknowledge the financial support provided by Fatima Jinnah Women University (FJWU), Pakistan through its research grant program.

## Notes

### Competing Interest Statement

The authors have declared no competing interest.

### Funding Statement

The study received the Funding for Experiment.

### Author Declarations

This study used only publicly available, anonymized datasets that do not require ethical approval.

